# Resilience and its determinants among adolescents and young adults with perinatally acquired HIV enrolled in a peer-led mentorship program in India

**DOI:** 10.64898/2026.04.08.26350433

**Authors:** Anita Shet, Michael Babu Raj, Siddha Sannigrahi, Babu Seenappa, Lakshmikanth Reddy, Ashley A. Sharma, Akshay G. Narayanan, SK Satish Kumar, Lakshmi Ganapathi

**Author notes:** Equal contribution (Anita Shet, Michael Babu Raj). **Statements and declarations**. **Competing interests** The authors declare that they have no competing interests, financial or non-financial, relevant to the content of this article. **Data availability** The datasets generated and analyzed during the current study are not publicly available due to the sensitive nature of the data involving a vulnerable adolescent population with HIV, but are available from the corresponding author upon reasonable request and subject to institutional data sharing agreements. **Author contributions** AS and MR conceptualized the study and designed the I'mPossible Fellowship intervention. AS, SS, AS2 and LG led data analysis and interpretation. BS, LR, and SKS contributed to data curation and fieldwork supervision. AS, LG and AGN oversaw the manuscript writing. All authors reviewed and approved the final manuscript.

## Abstract

**Background:** Adolescents and young adults with perinatally acquired HIV (APHIV) face complex psychosocial and structural challenges that may undermine resilience, a modifiable psychosocial determinant of treatment engagement, and health outcomes. Evidence on peer-led interventions targeting resilience among APHIV in South Asia remains limited. We evaluated resilience and its correlates among participants in the I’mPossible Fellowship, a peer-led mentorship intervention in India.

**Methods:** We conducted a cross-sectional evaluation of 216 APHIV following completion of the 24-month I’mPossible Fellowship in southern India in 2024. Surveys administered by trained youth investigators assessed sociodemographic, educational, and clinical characteristics.

Resilience was measured using the Child and Youth Resilience Measure-Revised (CYRM-R), a validated multidimensional tool capturing personal and relational resilience dimensions. Low resilience was defined as CYRM-R threshold score ≤33rd percentile. Multivariate logistic regression identified independent correlates of low resilience, and sensitivity analyses explored alternative CYRM-R thresholds.

**Results:** Participants had a mean age of 18.7 years (range 9-24); 50% had no surviving parents, and 43% lived in child care institutions. Median resilience scores were high (74, Interquartile range [IQR] 69-78), and 91% achieved viral suppression. In multivariate analyses, three factors were independently associated with low resilience: loss of both parents (adjusted odds ratio [aOR] 4.35, 95% CI 2.09–9.06), school discontinuation (aOR 2.43, 95% CI 1.10-5.34), and self-reported communication barriers at HIV clinics (aOR 5.83, 95% CI 2.69-12.64). These associations were consistent across sensitivity analyses at alternative resilience thresholds. At the most stringent threshold of low resilience (CYRM-R score ≤15th percentile), unsuppressed viral load also emerged as a significant correlate, suggesting that treatment failure may be concentrated among those with the most severely compromised resilience.

**Conclusions:** APHIV participating in a peer-led mentorship program demonstrated high overall resilience and viral suppression, but also revealed addressable vulnerabilities mapping to specific programmatic priorities. Peer-led models offer a promising foundational platform; however, complementary structural and psychosocial enhancements targeting these modifiable determinants are essential to optimize outcomes for those facing the greatest cumulative adversity.

## INTRODUCTION

Globally, nearly 1.57 million adolescents aged 10-19 were living with HIV in 2024, with the burden concentrated in Africa and Asia [1-3]. While antiretroviral therapy (ART) has substantially reduced HIV-attributable mortality [4–6], adolescents and young adults represent the sole age demographic experiencing a rise in HIV mortality [3,7]. Among those most at risk are adolescents and young adults with perinatally acquired HIV (APHIV), who navigate the compounding demands of chronic illness management, along with neurocognitive and psychosocial challenges [8]. This convergence substantially elevates risk for common mental health disorders, disengagement from HIV care, and poor virological outcomes [9–12]. During the transition to adult care, mortality among APHIV has been reported to be approximately tenfold higher than among HIV-negative peers [13], underscoring the urgency of psychosocial interventions tailored to this population.

Resilience, described as the capacity to adapt positively in the face of adversity, is a modifiable psychosocial determinant that mediates the relationship between structural disadvantage and health outcomes among APHIV. Strengthening resilience through targeted intervention has the potential to improve treatment engagement, adherence, and long-term wellbeing. Peer support models are particularly well-suited to this goal: grounded in the recognition that adolescence is a developmental stage of heightened peer sensitivity, these models leverage mentors with shared lived experience to provide emotional, informational, and practical support, promote positive coping, and normalize engagement with care [14,15]. Evidence from several countries in Africa, including the *Zvandiri* intervention in Zimbabwe, Project YES! in Zambia, and the *Boost* peer intervention in Malawi, Tanzania, Uganda, and Zambia, highlight the beneficial effects of peer support on retention in HIV care, stigma reduction, ART adherence, and psychosocial well-being among APHIV [16–20]. However, comparable evidence from Asia, including India, which carries the second highest HIV-affected population globally, remains limited [21,22]. India’s

National AIDS Control Program provides free ART nationally, yet challenges related to loss to follow-up, adverse outcomes, and mortality among APHIV persist [23–26].

Given this context, and the potential of peer support interventions to address the holistic treatment needs of APHIV, we developed the I’mPossible Fellowship, a peer-led mentorship intervention for youth with HIV, implemented in 2021 using community participatory action methods and grounded in the social ecological model (SEM) of health [27]. The SEM frames individual health as shaped by interacting determinants across interpersonal, institutional, and structural levels, a framework directly relevant to understanding why some APHIV remain resilient while others do not, and which levels of the system require targeted intervention. In this study, we evaluated resilience and its sociodemographic, educational, and clinical correlates among the first cohort of I’mPossible Fellowship participants, with the aim of identifying the specific subgroups and structural conditions for which peer mentorship may need to be augmented by complementary supports.

## METHODS

### The I’mPossible Fellowship intervention

The I’mPossible Fellowship intervention was implemented in the southern Indian states of Karnataka and Tamil Nadu in collaboration with government ministries with established HIV care initiatives [43]. Young adults with HIV (fellows) are selected based on their successful self-management, educational achievement, and demonstrated leadership, and undergo structured training based on the ICAP adolescent HIV care curriculum [39] to serve as mentors to younger APHIV (peers). Each fellow is matched with 20-25 peers by area of residence and mutual consent, with the fellowship period spanning 24 months. Fellow-peer contact comprises individual sessions (monthly, 20–30 minutes, by phone, video, or in-person) focused on health, psychosocial needs, and educational and career goals, and group sessions (quarterly, 60–90 minutes, in-person) addressing community-level issues and skill-building. Full details of session content and structure are provided in Table 1. Fellows provide two additional supports on behalf of the peers: health care navigation by liaiasing with ART center staff, and academic achievement and retention within the school system (Fig.1). Further fellows are also expected to maintaining their own health and continuing their own education

**Table 1:** Fellow interactive sessions in the I’mPossible Program intervention.

| Type of session | Channel of contact | Frequency and duration | Objectives | Examples of topics |
| --- | --- | --- | --- | --- |
| ***One-on-one sessions with peers*** | Phone, online video conferencing, or in-person | Every 4 weeks; 20-30 mins per session | To provide mentorship focused on individual peers' health, psychosocial and educational needs. | • Adherence and viral load<br>• Disclosure<br>• Navigating healthcare<br>• Stigma, coping mechanisms<br>• Counseling needs |
|  |  |  |  | • School and learning challenges |
| ***Group sessions with peers*** | In-person (~ 8 peers per group). | Every 3 months for each peer; 60-90 mins per group meeting | To discuss broader issues related to community needs, conduct skill-building sessions in communication | • HIV health<br>• Mental health<br>• Educational and life/career goals<br>• Gender issues<br>• Social relationships |

**Fig.1.**
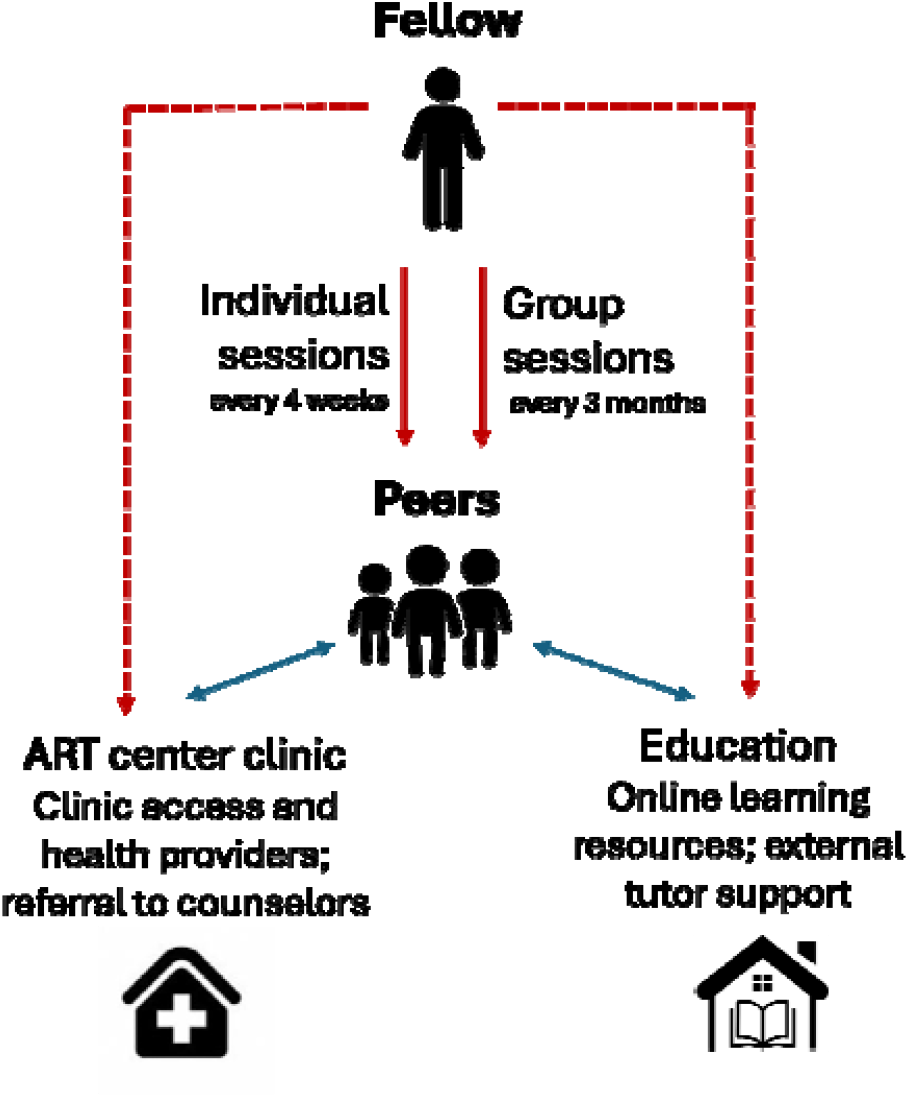
I’mPossible intervention diagram. Red arrows indicate intervention components, and blue arrows indicate existing relationships. Solid lines represent structured actions, dotted lines indicate unstructured ‘as needed’ actions

### Study population and data collection

The first I’mPossible Fellowship cohort enrolled 220 APHIV. For participants under 18 years, both caregiver/legal guardian consent and participant assent were obtained, while those ≥18 years provided direct consent. Following completion of the 24-month fellowship, 216 were available for the study (2 lost to follow-up, 2 declined participation). Between January and March 2024, cross-sectional data were collected. Incorporating principles of community-based participatory research [53], trained youth investigators administered surveys assessing sociodemographic, clinical, educational, and employment characteristics. Viral load from the past 12 months was obtained from participants’ ART cards.

Peer resilience was measured via the Child and Youth Resilience Measure-Revised (CYRM-R), a validated 17-item tool yielding a total score (range 17–85) across two subscales, personal resilience and relational resilience [28,29]. The CYRM-R tool is validated in diverse settings including India [30–32]. The CYRM-R was translated into Kannada, back-translated, and pilot-tested prior to deployment. Detectable viral load was defined as ≥150 copies/mL [33,34], and optimal ART adherence as fewer than 7 days of missed medication in the prior month. Low resilience was defined as a total CYRM-R score ≤71 (≤33rd percentile) [40,41].

Descriptive statistics summarized participant characteristics. Variables with prior evidence of association with resilience [35–37] were included in both univariate and multivariate logistic regression models estimating correlates of low resilience. Sensitivity analyses explored alternative low-resilience thresholds at the 15th, 25th, and 50th percentiles (scores ≤67, 69, and 74). All analyses were performed using SPSS version 29.0 (IBM Corp, Armonk, New York).

### Ethics

Ethical approval was obtained from the Institutional Review Boards of the Johns Hopkins Bloomberg School of Public Health (# 23077) and the Y.R. Gaitonde Centre for AIDS Research and Education (# YRG 375).

## RESULTS

### Peer characteristics

Among 216 APHIV peers, mean age was 18.7 years (range 9-24); 140 (64.8%) identified as male; 92 (42.6%) lived in or had recently graduated from a child care institution; and 108 (50.0%) had no surviving parents. (Table 2). Among 33.3% who had discountinued school, main reasons were financial constraints, parental loss and academic struggles; only 6.9% re-entered formal education during the fellowship period. Among the 38.4% currently employed, 62.2% reported workplace challenges including insufficient income, health concerns, and fear of HIV status disclosure. All peers were on ART, with the most common regimen being a single once-a-day pill of combined tenofovir/lamivudine/dolutagravir (TLD). Self-reported adherence to medications was high, with 98.1% reporting fewer than 7 missed doses in the prior month, and 91.4% achieved viral suppression (viral load <150 copies/mL). While 94.9% reported seeking help for physical symptoms, notably 28.2% reported being unable to discuss their problems at their HIV clinic, a finding that emerged as the strongest independent predictor of low resilience in subsequent analyses.

**Table 2.**
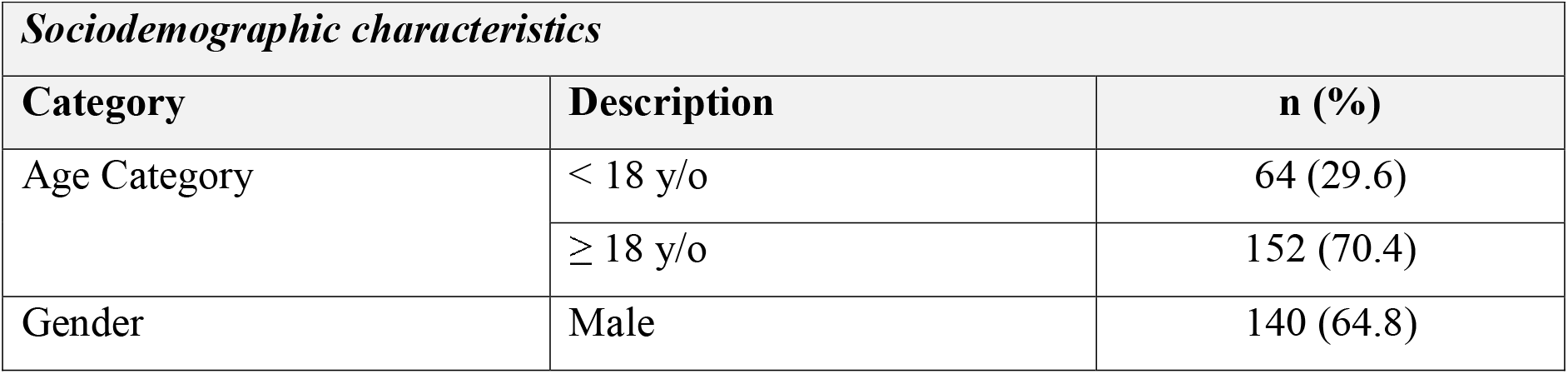
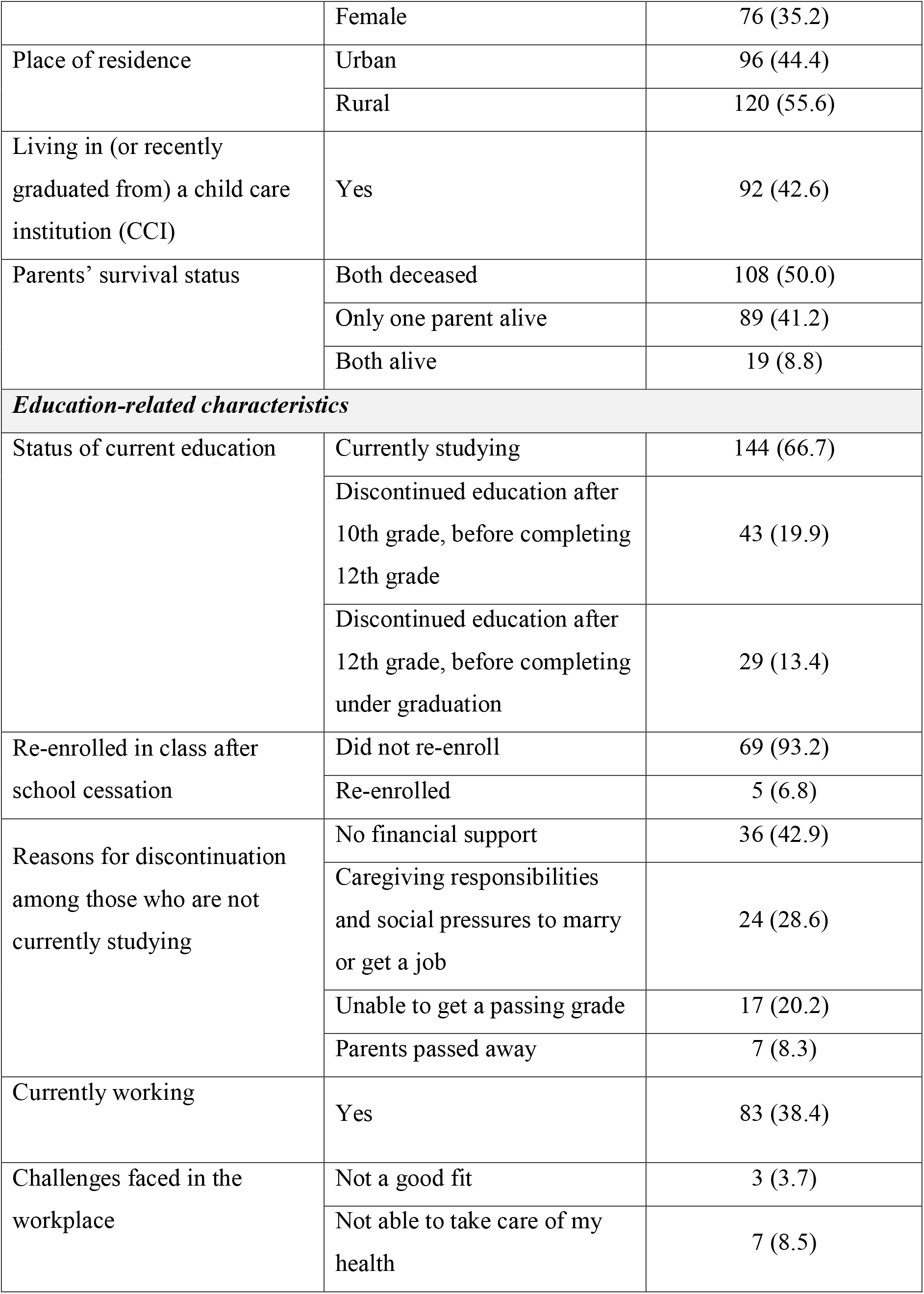
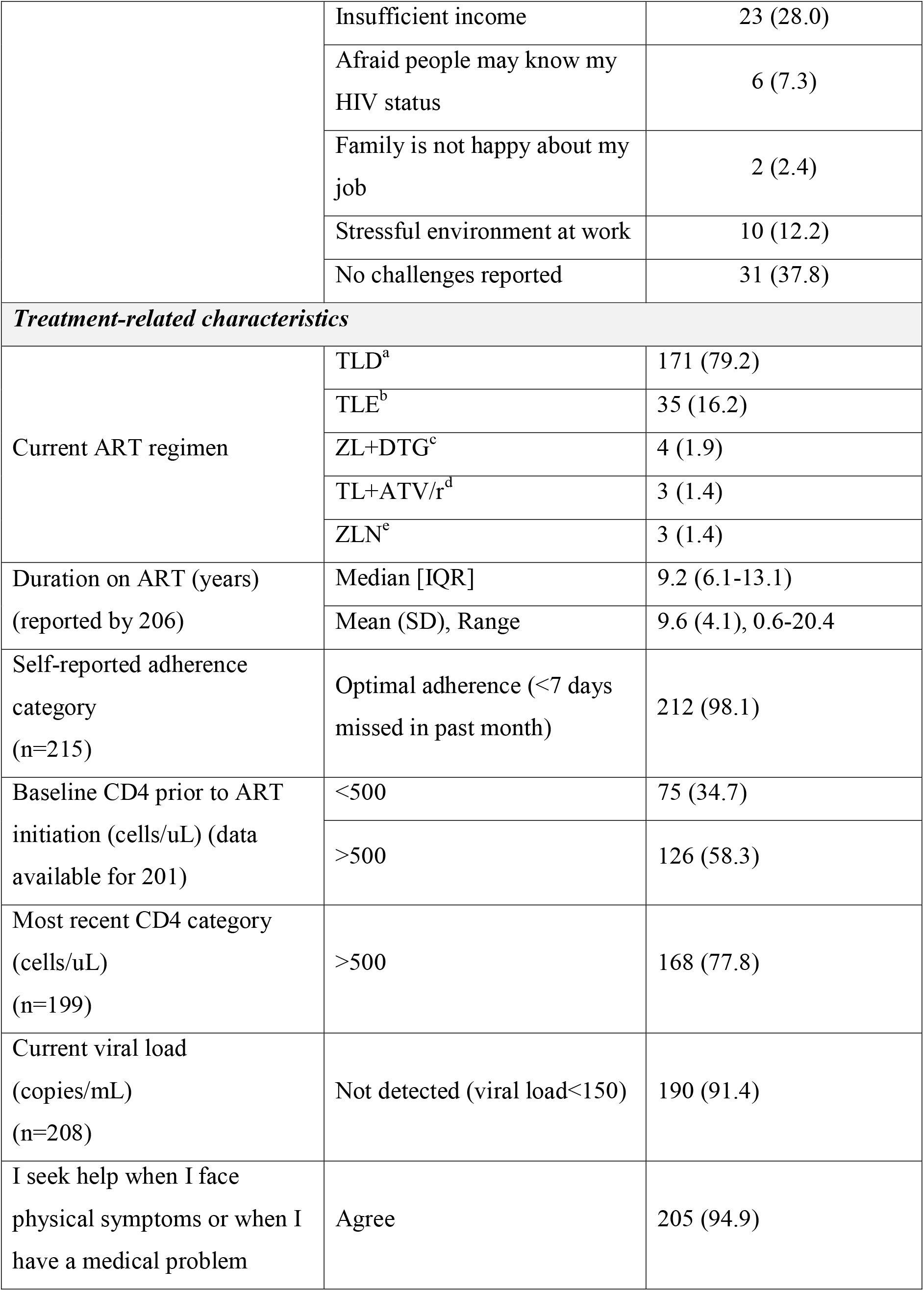
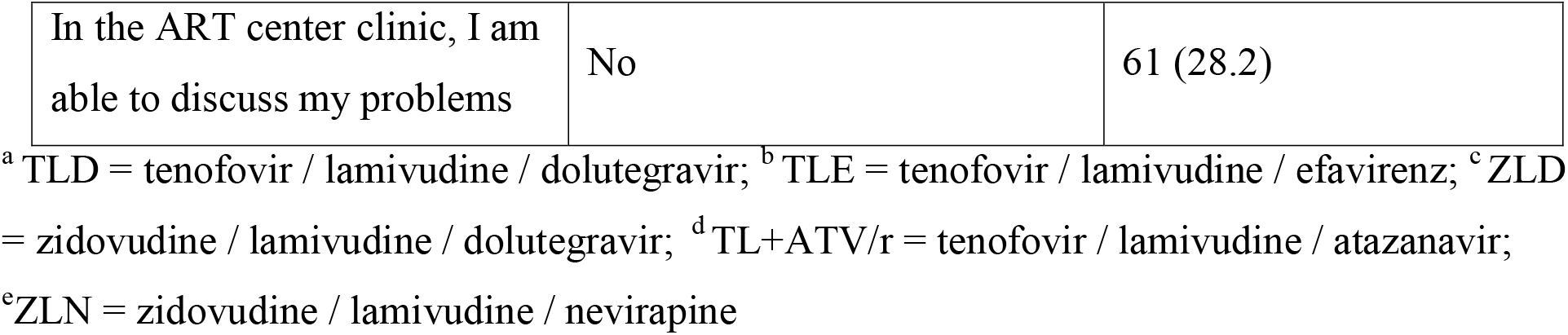
Characteristics of 216 APHIV peer participants in the I’mPossible fellowship program.

| ***Sociodemographic characteristics*** |  |  |
| --- | --- | --- |
| **Category** | **Description** | **n (%)** |
| Age Category | < 18 y/o | 64 (29.6) |
| | $\geq 18$ y/o | 152 (70.4) |
| Gender | Male | 140 (64.8) |
|  | Female | 76 (35.2) |
| Place of residence | Urban | 96 (44.4) |
|  | Rural | 120 (55.6) |
| Living in (or recently graduated from) a child care institution (CCI) | Yes | 92 (42.6) |
| Parents' survival status | Both deceased | 108 (50.0) |
|  | Only one parent alive | 89 (41.2) |
|  | Both alive | 19 (8.8) |
| ***Education-related characteristics*** |  |  |
| Status of current education | Currently studying | 144 (66.7) |
|  | Discontinued education after 10th grade, before completing 12th grade | 43 (19.9) |
|  | Discontinued education after 12th grade, before completing under graduation | 29 (13.4) |
| Re-enrolled in class after school cessation | Did not re-enroll | 69 (93.2) |
|  | Re-enrolled | 5 (6.8) |
| Reasons for discontinuation among those who are not currently studying | No financial support | 36 (42.9) |
|  | Caregiving responsibilities and social pressures to marry or get a job | 24 (28.6) |
|  | Unable to get a passing grade | 17 (20.2) |
|  | Parents passed away | 7 (8.3) |
| Currently working | Yes | 83 (38.4) |
| Challenges faced in the workplace | Not a good fit | 3 (3.7) |
|  | Not able to take care of my health | 7 (8.5) |
|  | Insufficient income | 23 (28.0) |
|  | Afraid people may know my HIV status | 6 (7.3) |
|  | Family is not happy about my job | 2 (2.4) |
|  | Stressful environment at work | 10 (12.2) |
|  | No challenges reported | 31 (37.8) |
| ***Treatment-related characteristics*** |  |  |
| Current ART regimen | TLD<sup>a</sup> | 171 (79.2) |
|  | TLE<sup>b</sup> | 35 (16.2) |
|  | ZL+DTG<sup>c</sup> | 4 (1.9) |
|  | TL+ATV/r<sup>d</sup> | 3 (1.4) |
|  | ZLN<sup>e</sup> | 3 (1.4) |
| Duration on ART (years)<br>(reported by 206) | Median [IQR] | 9.2 (6.1-13.1) |
|  | Mean (SD), Range | 9.6 (4.1), 0.6-20.4 |
| Self-reported adherence category<br>(n=215) | Optimal adherence (<7 days missed in past month) | 212 (98.1) |
| Baseline CD4 prior to ART initiation (cells/uL) (data available for 201) | <500 | 75 (34.7) |
|  | >500 | 126 (58.3) |
| Most recent CD4 category (cells/uL)<br>(n=199) | >500 | 168 (77.8) |
| Current viral load (copies/mL)<br>(n=208) | Not detected (viral load<150) | 190 (91.4) |
| I seek help when I face physical symptoms or when I have a medical problem | Agree | 205 (94.9) |
| In the ART center clinic, I am able to discuss my problems | No | 61 (28.2) |

<sup>a</sup> TLD = tenofovir / lamivudine / dolutegravir; <sup>b</sup> TLE = tenofovir / lamivudine / efavirenz; <sup>c</sup> ZLD = zidovudine / lamivudine / dolutegravir; <sup>d</sup> TL+ATV/r = tenofovir / lamivudine / atazanavir; <sup>e</sup> ZLN = zidovudine / lamivudine / nevirapine

### Resilience measures of peers

Median total CYRM-R score was 74 (IQR 69–78), with median personal and relational subscale scores of 43 (IQR 40–46) and 31 (IQR 28–33), respectively; 72 participants (33%) met the study definition of low resilience. In univariate analysis, loss of both parents, and school discontinuation, and inability to discuss problems at the ART clinic as well as longer duration on ART were significantly associated with low resilience. In multivariate analysis, three factors were independently associated with significantly greater odds of low resilience: inability to discuss problems at the ART clinic (aOR 5.83, 95% CI 2.69–12.64, p<0.001), loss of both parents (aOR 4.35, 95% CI 2.09–9.06, p<0.001), and school discontinuation (aOR 2.43, 95% CI 1.10–5.34, p=0.027) (Table 3). Age, gender, place of residence, living in a child care institution, current employment, and unsuppressed viral load were not significantly associated with low resilience in multivariate analysis. Sensitivity analyses across alternative low-resilience thresholds (≤15th, ≤25th, and ≤50th percentiles) confirmed that loss of both parents, school discontinuation, and clinical communication barriers consistently predicted greater odds of low resilience. Notably, at the most stringent threshold (≤15th percentile), unsuppressed viral load also emerged as a significant correlate, suggesting that treatment failure may be concentrated among those with the most severely compromised resilience.

**Table 3.**
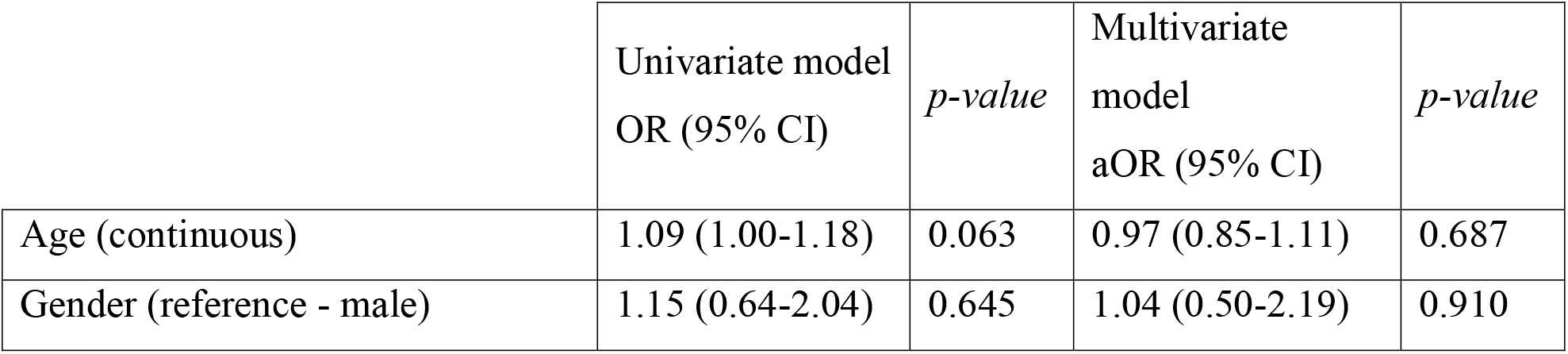
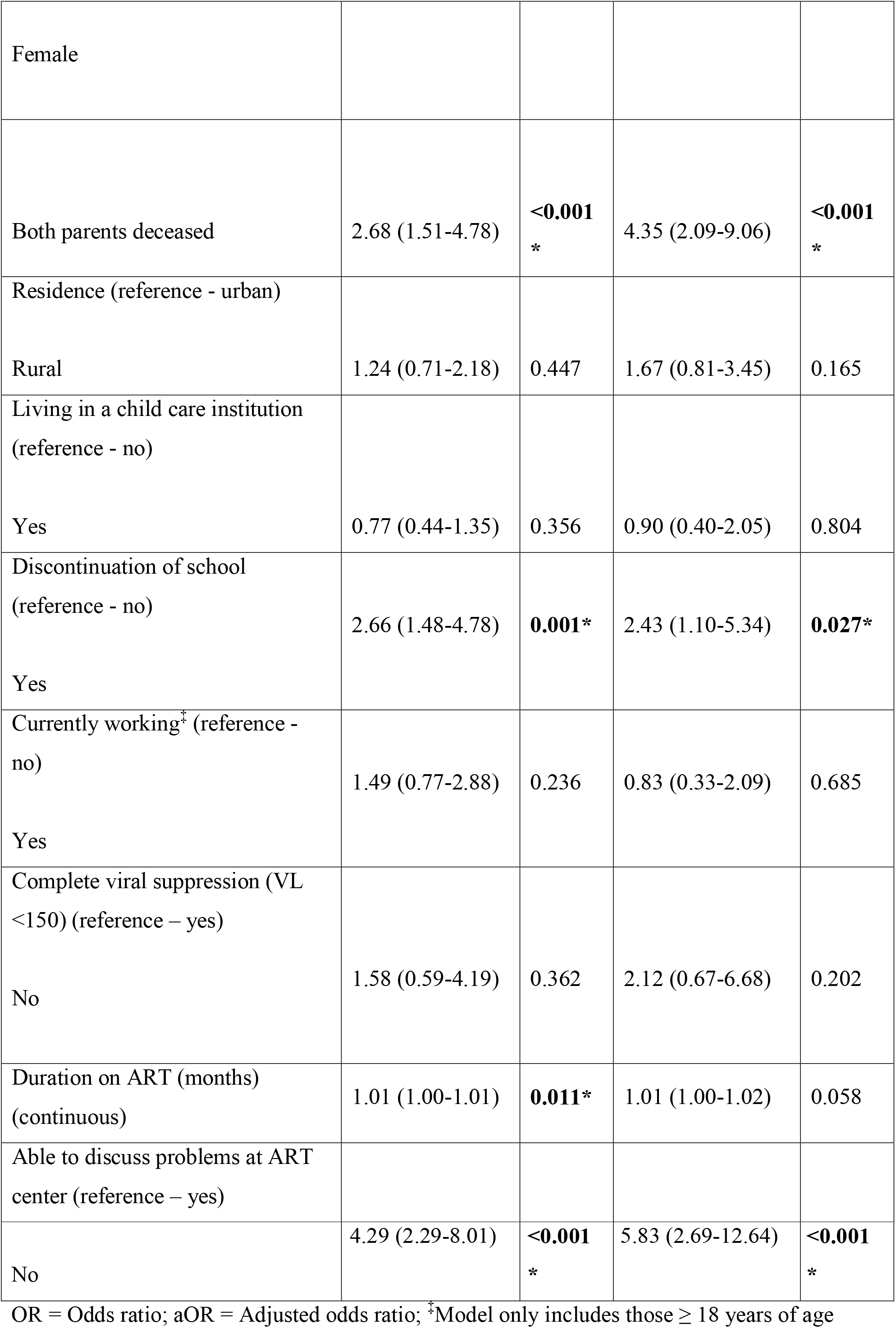
Correlates of low resilience (total CYRM-R score ≤33^rd^ percentile among 216 APHIV peers of the I’mPossible Fellowship program)

## DISCUSSION

This study presents early evidence from a peer-led mentorship intervention among APHIV in India, demonstrating high levels of viral suppression and resilience among participants in the I’mPossible Fellowship. While resilience scores were generally high, three discrete and independently significant correlates of low resilience emerged: loss of both parents, and school discontinuation, and communication barriers at HIV clinical settings. These patterns identify addressable leverage points where augmented scaffolding of multi-level supports within and alongside peer-led models is most likely to be needed to strengthen psychosocial and clinical outcomes.

Resilience is increasingly recognized as a dynamic, multisystemic process shaped by individual and socio-ecological factors rather than a fixed personal attribute [42]. Consistent with this conceptualization, our findings show that resilience was generally high among this cohort of APHIV but was influenced by relational and structural contexts. The social ecological model grounding the I’mPossible Fellowship which recognizes individual health as shaped by interpersonal, structural, and institutional determinants, maps directly onto this pattern: the three significant correlates of low resilience each operate at a different level of the socio-ecological system. Loss of both parents reflects relational and familial disruption; school discontinuation reflects structural and economic constraints; and communication barriers reflect institutional-level failures. The implication for program design is that peer mentorship, which operates primarily at the interpersonal level, will need additional targeted components to fully address vulnerabilities operating at these other levels.

Loss of both parents emerged as one of the strongest correlates of low resilience, reflecting the central role of caregiver relationships in buffering adversity and promoting psychological adaptation. Youth who experience parental loss in early childhood often face not a single traumatic event but the ongoing absence of primary attachment figures, compounded by financial instability and reduced social support, all of which disrupt the protective environments that sustain adaptive coping and positive development [45]. This finding is consistent with our prior work, in which loss of both parents was significantly associated with increased odds of anxiety among this same population [46], suggesting that the psychosocial consequences of caregiver loss extend across multiple domains of wellbeing. Notably, living arrangement (child care institution versus community) was not associated with resilience differences, suggesting that institutional care does not itself confer protection or harm against these relational losses; the vulnerability travels with the youth regardless of where they live. This has direct programmatic implications: grief-informed and trauma-responsive services are needed as enhancements within peer mentorship models specifically for bereaved youth, and linkage to social protection schemes addressing the material consequences of caregiver loss is an important complement.

School engagement is frequently protective for adolescents, providing not only educational opportunity but also daily structure, social connectedness, and identity formation [36]. A study from Malawi showed that school attendance is associated with lower odds of depression among adolescents with HIV [44], and while our outcome is resilience rather than depression, the conceptual overlap is relevant. In our cohort, educational discontinuation was independently associated with low resilience, and re-entry during the fellowship was modest, suggesting that that once broken, educational pathways are difficult to restore. The most commonly cited barrier to continuation was financial, and nearly all peers (93.2%) identified financial support as a critical need. These are material barriers that peer mentorship alone cannot address. While the I’mPossible fellowship actively supports peers’ academic and career goals, these findings point to the need for structured linkage to scholarship programs, tuition assistance, and social protection schemes as formal components of intervention design, rather than supplementary add-ons. Supporting educational retention before discontinuation occurs, rather than attempting re-entry after the fact, should be a prevention priority.

The strongest predictor of low resilience in this study was the inability to discuss problems at HIV clinic, a finding that warrants particular attention from both a programmatic and a health systems perspective. Communication barriers within clinical settings likely reflect a combination of stigma, power imbalances between youth and adult providers, insufficient confidentiality protections, and the absence of youth-friendly care models [38]. These barriers not only undermine psychosocial wellbeing directly, but plausibly disrupt ART adherence and engagement in care, the precise pathways through which resilience and treatment outcomes intersect. This is supported by our sensitivity analyses, in which unsuppressed viral load emerged as a significant correlate of low resilience at the most stringent threshold (≤15th percentile of CYRM-R scores). This pattern is consistent with evidence that depression and anxiety are associated with poorer ART adherence and viral suppression, and that addressing psychological distress can improve HIV outcomes [48,49,51,52]. In practical terms, it suggests an interdependent relationship: reduced resilience may compromise adherence and engagement with care, while poor treatment outcomes in turn exacerbate stress and further erode self-efficacy.

These findings make a strong case for integrating mental health screening and psychosocial support directly into HIV clinical encounters, particularly for youth with unsuppressed viral loads, and for redesigning ART clinic interactions to be youth-friendly.

The peer-led model of the I’mPossible Fellowship addresses multilevel determinants of resilience through a social ecological framework. The peer-led model provides informational, emotional, and affirmational support that strengthens adaptive coping, self-efficacy, and social connectedness among APHIV. Fellows who are young people with their own lived experience of HIV, serve as credible role models who promote health literacy, facilitate navigation of care systems, normalize help-seeking behavior, and reduce internalized stigma. Through structured mentorship grounded in shared identity, the fellowship reinforces both the personal and relational dimensions of the resilience construct, and the high overall resilience and viral suppression rates in this cohort are consistent with peer mentorship contributing meaningfully to these outcomes (Fig. 2).

**Fig.2.**
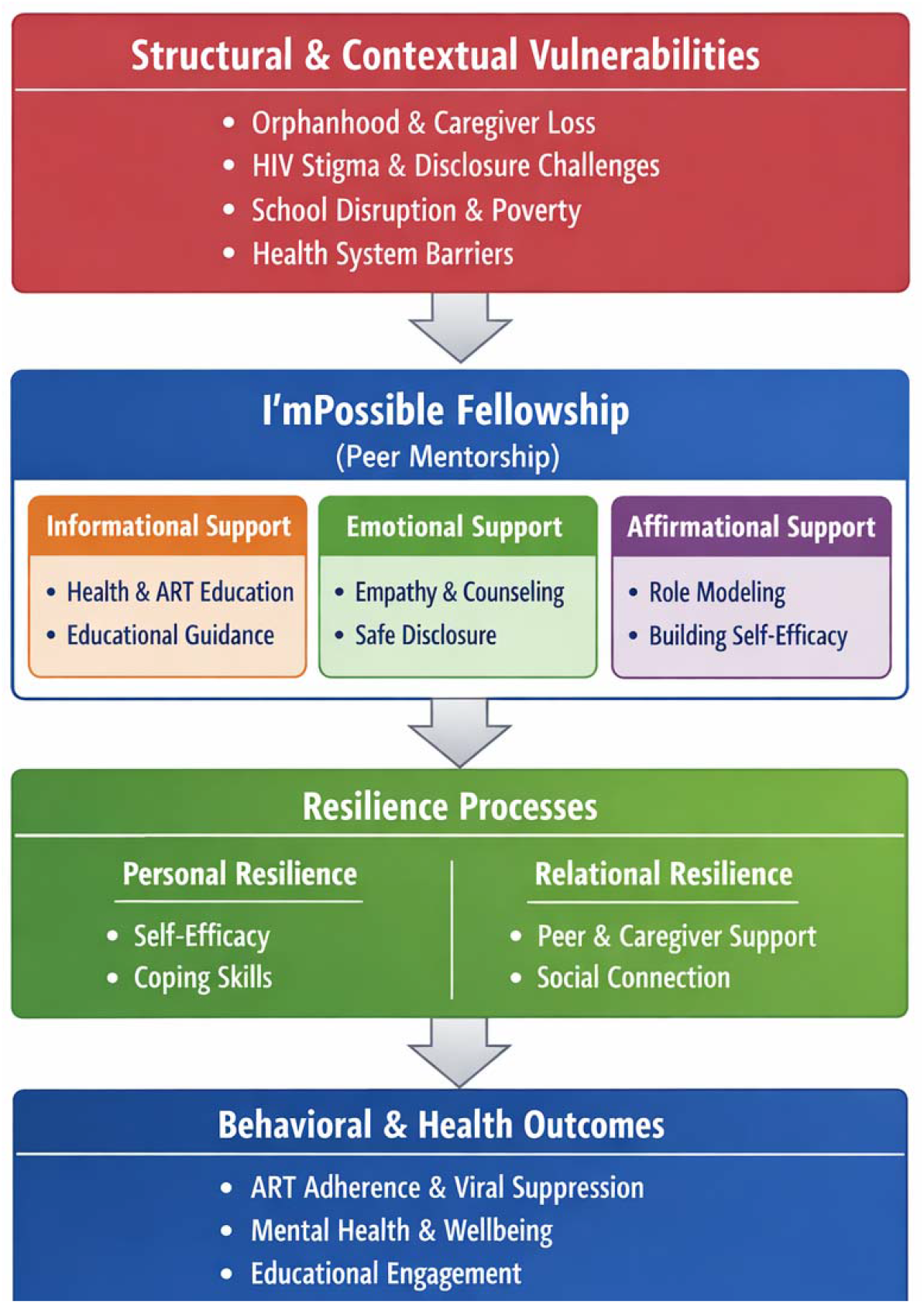
Conceptual framework of resilience among adolescents and young adults with perinatally acquired HIV (APHIV) within the I’mPossible Fellowship. The model illustrates how structural vulnerabilities shape resilience, and how peer mentorship provides informational, emotional, and affirmational support that strengthens personal and relational resilience, leading to improved psychosocial and HIV-related outcomes

Evidence from comparable peer-led programs in sub-Saharan Africa including Zimbabwe’s Zvandiri program and Zambia’s Project YES! similarly demonstrates improvements in viral suppression and psychosocial wellbeing after integrating peer mentors into HIV care settings [19,20,47]. Evidence from Asia, drawn from peer education/outreach models is similarly suggestive of programmatic success with adolescents and young adults at risk for HIV [50]. Qualitative evidence from our own evaluation of the I’mPossible Fellowship found that fellows were seen as role models who created safe spaces for peers to discuss challenges without fear of stigma, providing informational, emotional, and affirmational support that enhanced resilience [43]. As fellows were also young people with lived experience of HIV, they were uniquely positioned to provide credible role modeling, emotional validation, and practical guidance to peers navigating similar challenges. This combination of shared identity and structured mentorship may have helped reduce isolation, normalize engagement with care, and facilitate adaptive coping strategies among peers thereby building resilience. The current findings extend this picture by identifying the specific subgroups and structural conditions of heightened vulnerablities where peer-led programming serves as a foundation for additional targeted and complementary enhancements.

This study’s strengths include a near-complete cohort follow-up rate (216 of 220 enrolled peers), the use of a validated, multidimensional resilience measure with established applicability in Indian settings, and the involvement of trained youth investigators in data collection. Several limitations warrant consideration. The cross-sectional design precludes causal inference between the intervention and resilience outcomes, and the absence of a comparison group limits attribution of observed outcomes to the intervention specifically. Self-reported measures may be subject to social desirability or recall bias. Critically, the study sample is drawn from youth already enrolled in a structured peer support program, meaning that APHIV not connected to such programs, and quite possibly a more vulnerable group, are entirely unrepresented. Findings may therefore underestimate the burden of low resilience in the broader APHIV population in India, and caution is warranted in generalizing results to youth outside comparable care structures.

## CONCLUSION

In summary, the three correlates of low resilience we identified in our analyses, (loss of both parents, educational discontinuation, and communication barriers in clinical settings), each point to specific, actionable enhancements: trauma-informed and grief-responsive psychosocial services with social protection linkage for bereaved youth; proactive educational retention strategies with financial assistance and vocational pathways; and youth-friendly ART service redesign with integrated mental health support. Future research should evaluate the longitudinal impact of enhanced peer-led mentorship on resilience trajectories and HIV care engagement that address specific vulnerabilities within youth-centered HIV service models across India and comparable settings.

## Data Availability

The datasets generated and analyzed during the current study are not publicly available due to the sensitive nature of the data involving a vulnerable adolescent population with HIV, but are available from the corresponding author upon reasonable request and subject to institutional data sharing agreements.

## Acknowledgements

We thank the adolescents and young adults who participated in this study and the youth investigators who co-led the research. We are grateful to the staff and community health workers at YRGCARE, Chennai, and RISHI Foundation, Bengaluru, for their support in intervention delivery and data collection.

